# SARS-CoV-2 variants in the making: Sequential intrahost evolution and forward transmissions in the context of persistent infections

**DOI:** 10.1101/2022.05.25.22275533

**Authors:** Ana S. Gonzalez-Reiche, Hala Alshammary, Sarah Schaefer, Gopi Patel, Jose Polanco, Juan Manuel Carreño, Angela A. Amoako, Aria Rooker, Christian Cognigni, Daniel Floda, Adriana van de Guchte, Zain Khalil, Keith Farrugia, Nima Assad, Jian Zhang, Bremy Alburquerque, PARIS/PSP study group, Levy Sominsky, Charles Gleason, Komal Srivastava, Robert Sebra, Juan David Ramirez, Radhika Banu, Paras Shrestha, Florian Krammer, Alberto Paniz-Mondolfi, Emilia Mia Sordillo, Viviana Simon, Harm van Bakel

## Abstract

Persistent severe acute respiratory syndrome coronavirus 2 (SARS-CoV-2) infections have been reported in immune-compromised individuals and people undergoing immune-modulatory treatments. Although intrahost evolution has been documented, to our knowledge, no direct evidence of subsequent transmission and stepwise adaptation is available.

Here we describe sequential persistent SARS-CoV-2 infections in three individuals that led to the emergence, forward transmission, and continued evolution of a new Omicron sublineage, BA.1.23, over an eight-month period. The initially transmitted BA.1.23 variant encoded seven additional amino acid substitutions within the spike protein (E96D, R346T, L455W, K458M, A484V, H681R, A688V), and displayed substantial resistance to neutralization by sera from boosted and/or Omicron BA.1-infected study participants. Subsequent continued BA.1.23 replication resulted in additional substitutions in the spike protein (S254F, N448S, F456L, M458K, F981L, S982L) as well as in five other virus proteins.

Our findings demonstrate that the Omicron BA.1 lineage can diverge further from its already exceptionally mutated genome during persistent infection in more than one host, and also document ongoing transmission of these novel variants. There is an urgent need to implement strategies to prevent prolonged SARS-CoV-2 replication and to limit the spread of newly emerging, neutralization-resistant variants in vulnerable patients.

## Introduction

The genomic landscape of severe acute respiratory syndrome coronavirus 2 (SARS-CoV-2) has been shaped by the emergence of antigenically diverse variants of concern (VOC)^1^. These viral variants carry mutations that render them more transmissible, more fusogenic, and/or permit escape not only from infection but also vaccine-induced immunity^2-4^. Some variants such as the Omicron VOCs, which swept the globe starting November 2021, also display partial or complete resistance to therapeutic or prophylactic monoclonal treatments^5^. Over the past ten months Omicron sublineages with increasing numbers of mutations in spike have continued to emerge (e.g., BA.1, BA.2, BA.4/5), posing great challenges to the containment of SARS-CoV-2 spread and providing the rationale for formulation of bivalent SARS-CoV-2 booster vaccines that include an Omicron BA.1 or BA.5 spike in addition to the ancestral spike.

Most patients with coronavirus disease 2019 (COVID-19) clear the virus upon resolution of the acute infection. However, ongoing SARS-CoV-2 replication has been documented in a subset of immunocompromised individuals. In these chronically infected cases, recovery of infectious virus over several months and stepwise acquisition of mutations in spike has been observed, pointing to positive selection^6-13^. It has been speculated that prolonged intrahost viral evolution played a role in the emergence of several SARS-CoV-2 VOCs such as Alpha and Omicron^14,15^, but clear evidence for forward transmissions of mutated variants from chronic infection cases has been lacking.

We describe herein a primary case of persistent SARS-CoV-2 Omicron BA.1 infection marked by intrahost evolution of a variant (Omicron BA.1.23) encoding seven additional amino acid substitutions in the already antigenically distinct Omicron BA.1 spike protein, and that resulted in at least five further cases of Omicron BA.1.23 infection. Persistent infections documented in two of these cases (one lasting four weeks, the other more than four months) were associated with continued BA.1.23 evolution and acquisition of additional mutations within and outside of spike. Although the variants that evolved in the persistently infected cases during the cumulative eight-month period reveal unique constellations of mutations in each, they also strongly point to convergent viral evolution with other co-circulating SARS-CoV-2 lineages. Notably, most amino acid changes occurred at positions known to confer either immune escape^16,17^ or altered viral fusogenicity^18-20^. Some of these escape mutations in BA.1.23 are now signature substitutions in emerging Omicron lineages such as BA.2.75.2, indicating that persistent viral replication in the context of suboptimal immune responses is an important driver of SARS-CoV-2 diversification.

## Results

### Emergence of a novel BA.1 sublineage through intrahost evolution

We performed genomic analysis of serially collected nasopharyngeal (NP) and anterior nares (AN) samples from an immunocompromised patient (P1) with diffuse B-cell lymphoma and persistent SARS-CoV-2 Omicron BA.1 replication between December 2021 and March 2022. Over a 12-week period, we documented the accumulation of nine amino acid substitutions in the spike protein N-terminal domain (NTD), the receptor binding domain (RBD), and in the S1/S2 furin cleavage site (FCS) (**Fig. 1**) within the same patient. The first four mutations R346T, K458M, E484V, and A688V were detected simultaneously 40 days after the initial SARS-CoV-2 diagnosis and were fixed. Two weeks later (day 64), L167T and the FCS mutation P681Y were detected in addition to the four initial mutations. During the following weeks (day 72 and day 81), two different viral populations emerged each carrying shared (L455W) as well as additional distinct signature mutations (E96D on day 72, S477D on day 81). All but one of the mutations observed in the consensus sequences outside the spike gene were non-synonymous, providing further support for positive intrahost selection of spike protein changes with competitive replication advantages.

**Figure 1.**
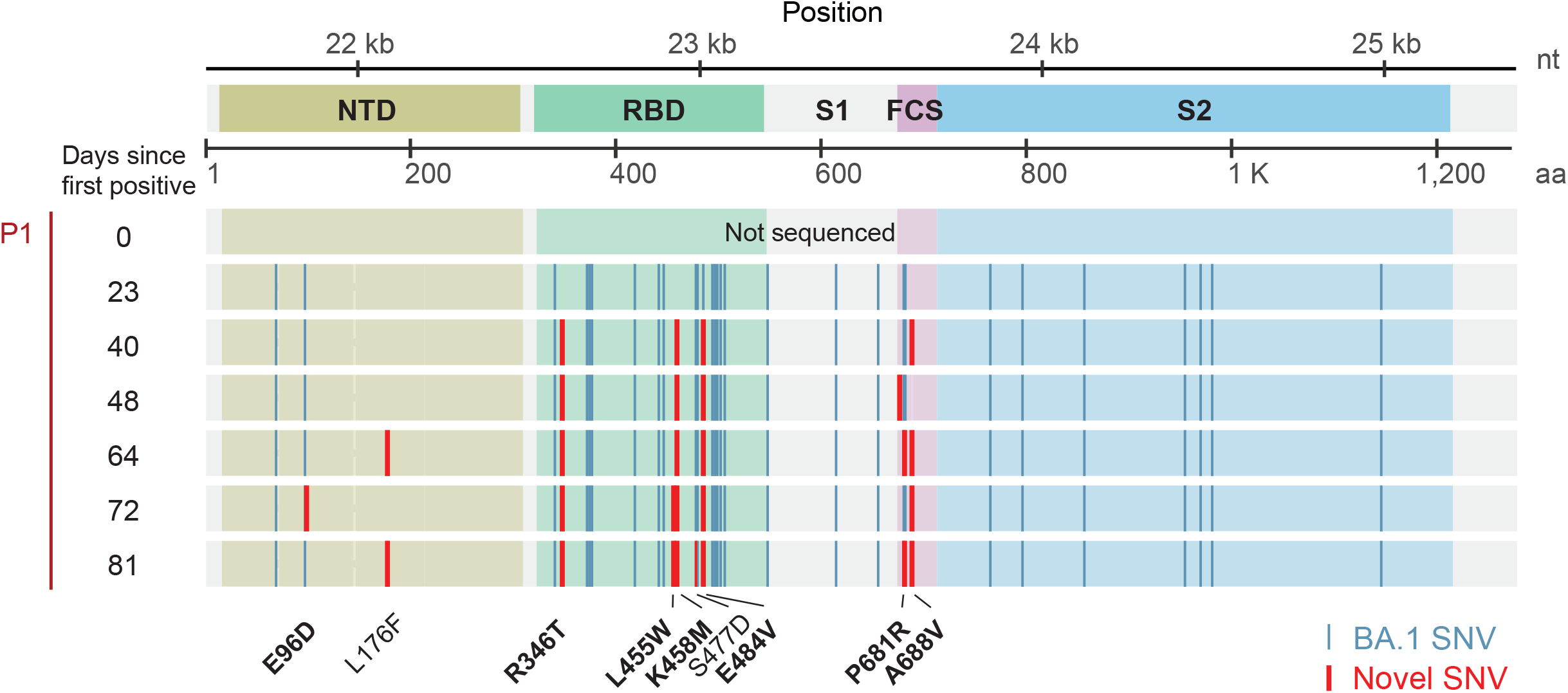
Intrahost emergence of an Omicron BA.1.23 subvariant. Multiple sequence alignment of the SARS-CoV-2 spike protein showing the appearance of nucleotide and amino acid substitutions in the spike protein in sequential specimens obtained from the index case (Patient 1, P1).

### Forward spread confirms transmission potential of the novel BA.1.23 sublineage

Background health system-wide SARS-CoV-2 genomic surveillance conducted by the Mount Sinai Pathogen Surveillance Program (MS-PSP) during the same time period identified three other patients harboring SARS-CoV-2 Omicron variants that shared seven of the amino acid substitutions found in the index case P1 (E96D, R346T, L455W, K458M, A484V, H681R, A688V), and were consistent with multiple forward transmissions based on the timeline of infection (**Fig. 2, Table 1**). Although viral isolation failed for specimens available from the index case, we successfully cultured SARS-CoV-2 from the initial positive specimens of the subsequent cases, confirming the transmission of replication-competent virus. All three forward transmissions were detected in patients with underlying hematologic malignancies.

**Table 1:**
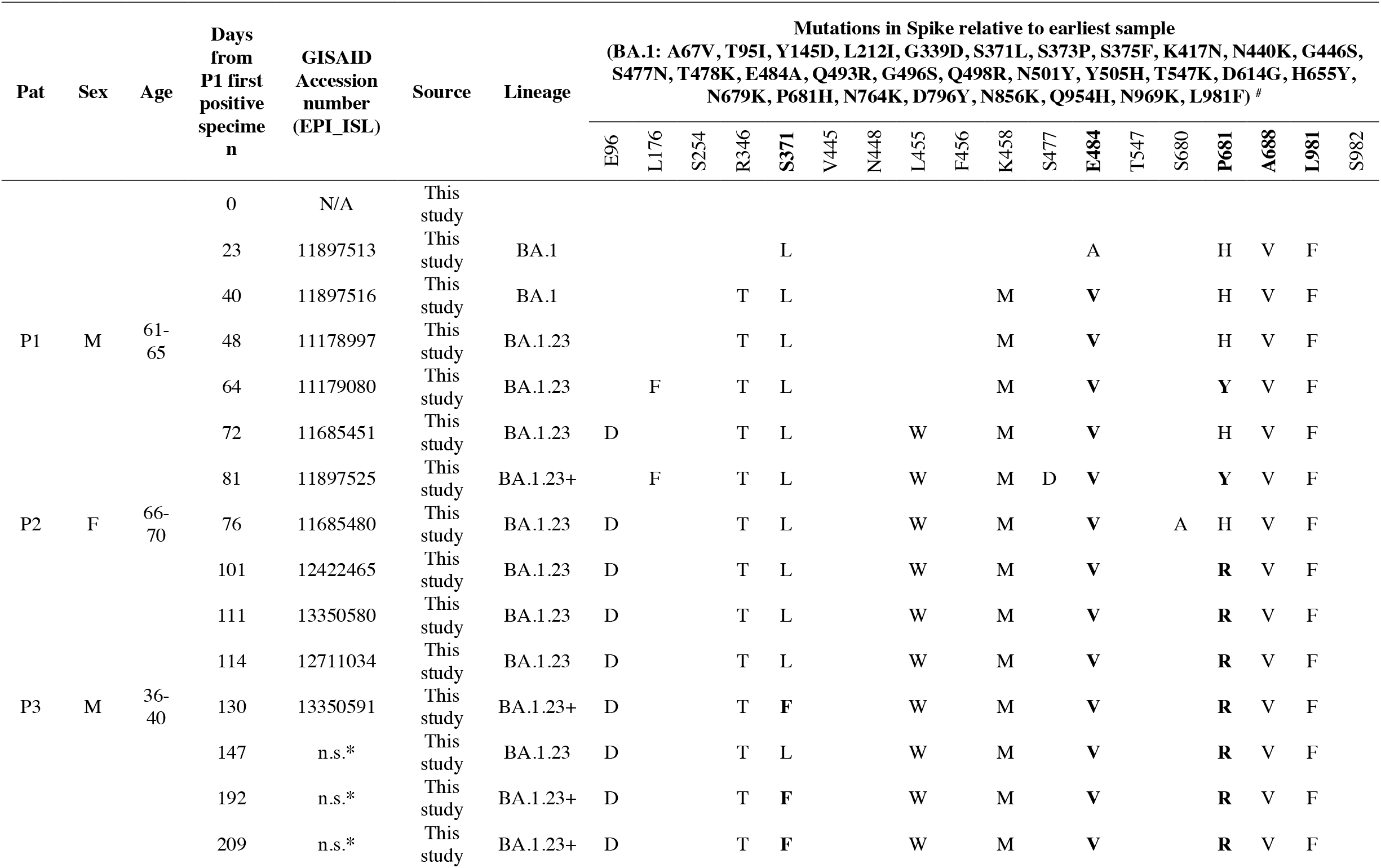
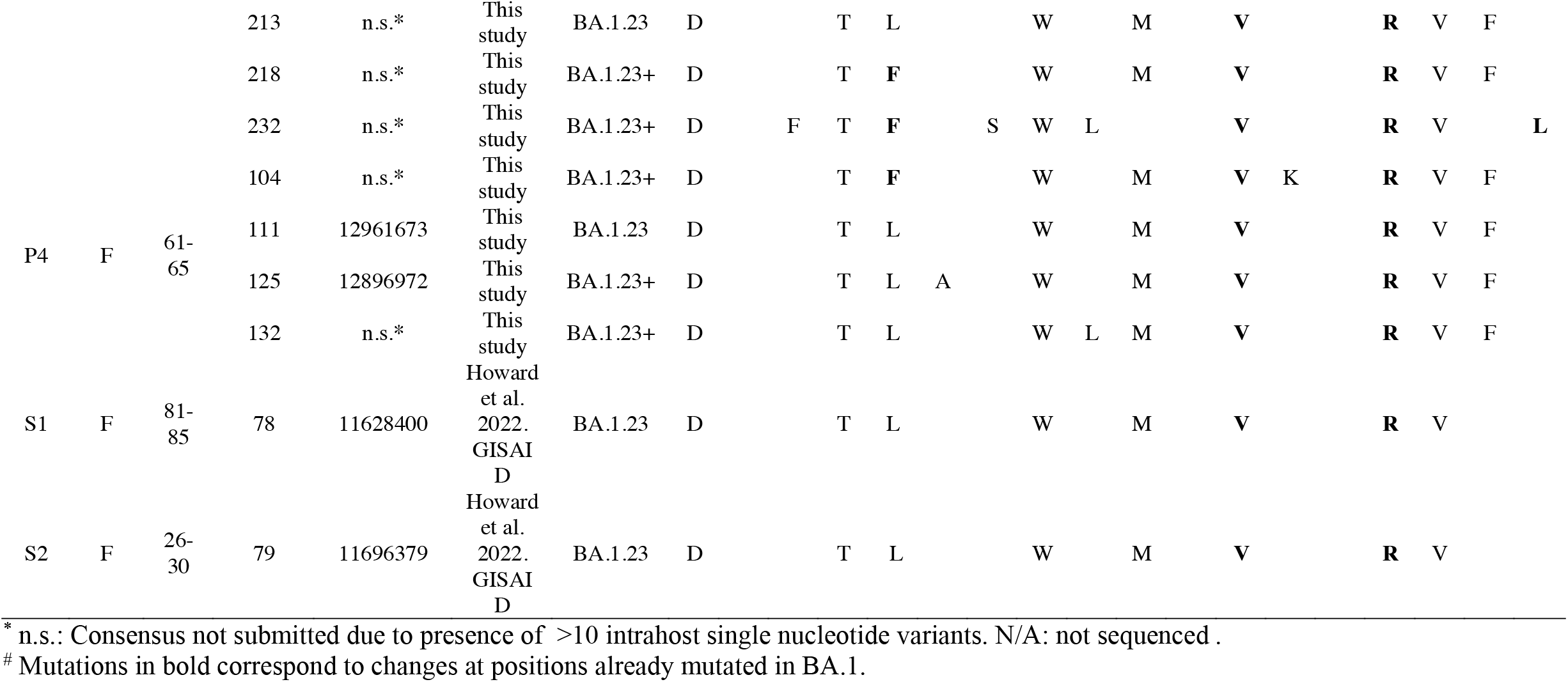
Spike mutations detected in the sequenced cases.

**Figure 2.**
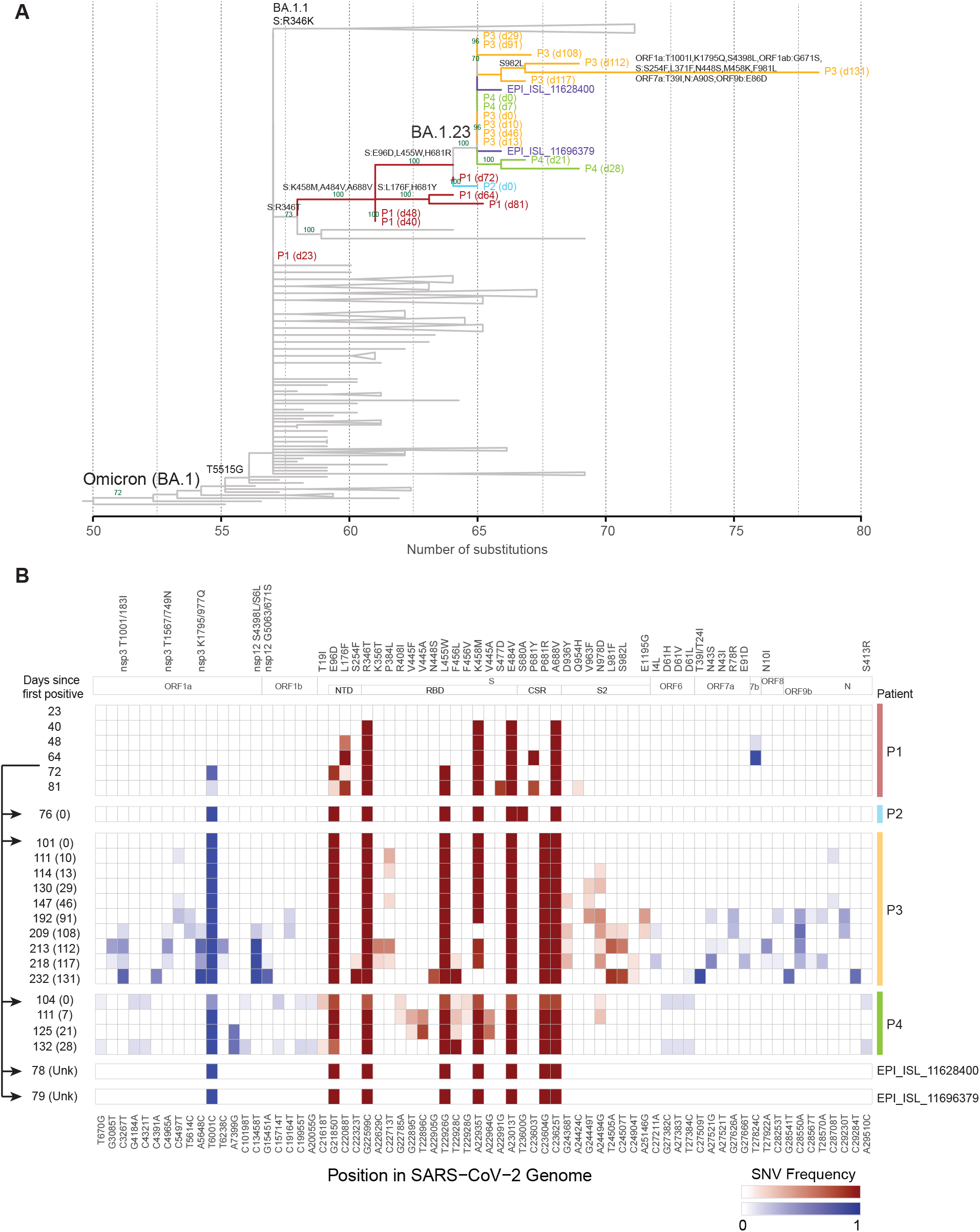
Forward transmission of the Omicron BA.1.23 subvariant. **A)** Maximum likelihood phylogenetic subtree with SARS-CoV-2 (BA.1) sequences from the persistent infection case P1 and the onward transmissions (P2, P3, P4), in a global background of sequences available in GISAID. Branches are colored to identify the patient. The number of days after the first SARS-CoV-2 positive specimen of P1 is indicated in brackets. Sibling clusters of BA.1 and other sublineages are collapsed for easier visualization. The x-axis shows the number of nucleotide substitutions relative to the root of the phylogenetic tree. Bootstrap support values above 70% are shown for the un-collapsed branches. The distinct BA.1 subvariant that was transmitted was designated as PANGO lineage BA.1.23. **B)** Frequency of single nucleotide variants (SNV), and amino acid substitutions for SARS-CoV-2 genomic positions with consensus changes from ancestral Wuhan-1 and Omicron BA.1. Positions with mixed nucleotides below consensus are also shown for intrahost SNVs (mSNVs) seen in more than one time point. The sequenced specimens are shown sequentially for patient 1 (P1) with prolonged BA.1 infection and transmission cases of BA.1.23 (P2, P3 and P4). The viral genomes from P1 show progressive accumulation of mutations in the N-terminal domain (NTD), receptor binding domain (RBD), and S1/S2 furin cleavage site (FCS); The same constellation of mutations was subsequently detected in three documented transmission cases (P2, P3 and P4). The number of days since the first positive test in P1 is shown on the left, with the number of days after the first positive test for each patient between brackets. The mutation frequencies within the Spike protein are shown in red, and for non-Spike genome regions in blue. Positions are numbered according to the reference genome sequence NC_045512.2. Arrows on the left indicate the likely window of transmission from the index patient between days 64-72.

Lastly, two additional related SARS-CoV-2 genomes from the NYC area were identified from the GISAID (Global Initiative on Sharing Avian Influenza Data) database (**Fig. 2**). Based on the metadata provided, these two cases differed by age and gender from our cases, consistent with independent but limited community spread of this new Omicron BA.1 sub-variant. All transmission cases contained one synonymous change in Orf1a (T6001C) previously only observed at day 72 in the index case (**Extended Data Fig. 1**), narrowing the time window of the first transmission to a few weeks following day 64. As of August 2022, the most recent case was detected in mid-April 2022 and the last SARS-CoV-2 positive specimen from a forward transmission case (P3) was collected in September 2022. Taken together, the presence of a unique combination of mutations in all six cases is indicative of forward transmission of this novel Omicron subvariant, which received the BA.1.23 Pango lineage designation.

### Sequential persistent infections drive further intrahost evolution of BA.1.23

While patient P2 cleared the BA.1.23 infection, patients P3 and P4 both developed persistent infections (**Fig. 2 and 3A**). The MS-PSP continued monitoring these patients during follow-up visits and hospitalizations. Patient P4 remained positive by nucleic acid amplification tests (NAAT) for four weeks, with acquisition of an additional mutation in the spike RBD (S:V445A) within one week of the initial positive test (**Fig. 2 and 3A**). Patient P3 developed a much longer persistent infection lasting for more than four months. Genomic analysis of all 10 serially collected specimens from P3 identified 14 additional amino acid substitutions throughout the viral genome (**Fig. 2B and Extended Data Fig. 1**). The most recent specimen collected 131 days after P3’s COVID-19 onset contained 11 additional amino acid substitutions compared to the originally transmitted BA.1.23 variant. These included six substitutions in the spike NTD (S254F), RBD (N448S, F456L, reversion of 458M to K), and S2 (reversion of 981L to F, S982L); as well as five substitutions in other SARS-CoV-2 proteins (ORF1a nsp3: T1001/183I, K1795/977Q, ORF1ab_nsp11 S4398/6L; ORF1ab_nsp12: G5063/671S; and ORF7a: T39I/T24I). Notably, two spike amino acid reversions to the WA1/USA (S:M458K) or BA.1 (S:F981L) state were accompanied by changes at neighboring positions (S:F456L and S:S982L) (**Fig. 2B and Extended Data Fig. 1**).

**Figure 3.**
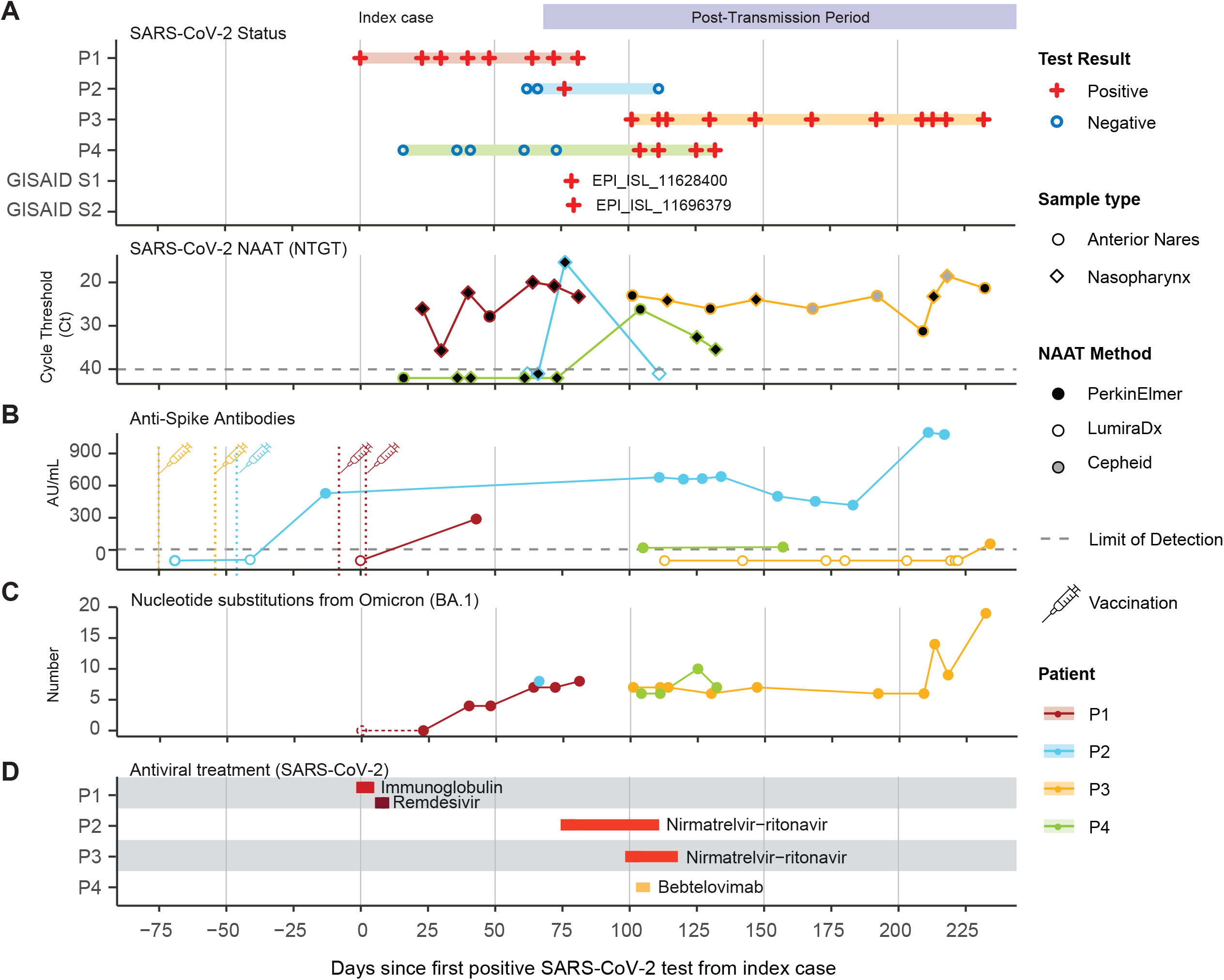
Timeline of BA.1.23 evolution compared to antibody levels and treatments. **A)** The timeline of BA.1.23 infected patients (top) is shown together with nucleic acid amplification test (NAAT) SARS-CoV-2 (N gene) cycle threshold (Ct) values for respiratory specimens, for different diagnostic methods (bottom). Empty markers indicate negative tests by alternative diagnostic tests with no Ct value readout (LumiraDxTM). **B)** Number of nucleotide substitutions in the consensus sequence relative to Omicron BA.1 in the sequenced specimens. **C)** SARS-CoV-2 spike binding IgG antibody levels for P1-P4. Antibody levels are shown in arbitrary units per mL (AU/mL). Available vaccine information is indicated. **D)** Timeline of SARS-CoV-2 antiviral treatments received by P1-P4. Treatment duration is indicated by the length of the bar.

We further identified multiple positions with intra-host single nucleotide variants that were present in only a minority of the SARS-CoV-2 viral populations, hereafter referred to as minor single nucleotide variants (mSNVs), within each of the three persistent infection cases (P1, P3, and P4). In some cases, we detected mSNVs within or outside the spike gene in earlier specimens, prior to their fixation in subsequent specimens from the same patient (**Figure 2B and Extended Data Figure 1,2**). We also observed numerous positions with mSNVs that persisted across multiple sequential specimens without becoming dominant. This was most notable for patient P3: between the first specimen collected on day 101 and the ninth specimen collected on day 218, the number of positions with mSNVs at a level >10% in two or more specimens increased gradually from 0 to 19 – eventually outnumbering the consensus sequence changes by a factor of three (**Fig. 2B and Extended Data Fig. 3**). Approximately half of these mutations occurred within the spike gene, where nonsynonymous mSNVs were clustered in the S1 (NTD:S254F, RBD:K356T, S371F, P384L, F456L, K458M, and FCS:N679K) and S2 (D936Y,V963F, N978D, L981F, S982L, E1195G) domains (**Fig. B2 and Extended Data Fig. 4**). Mutations at these positions are rare at the consensus level, with a maximum prevalence of 0.1% in the GISAID reference database [as of 2022-09-10]. The final specimen collected two weeks later, on day 232, showed eleven additional substitutions (including five at positions with prior mSNVs) and two positions with mSNVs (**Fig. 2B, Extended Data Fig. 3 and Supplementary Table S1**). The progressive accumulation of mutations as well as the low prevalence of the specific substitutions in co-circulating SARS-CoV-2 lineages in NYC at the time, makes it unlikely that the mutated genotypes are the result of recombination with other viral variants. The large shift in mutational profile may reflect a fitness advantage of the dominant variant in this specimen. Thus, intra-host adaptation occurs with different dynamics pointing to bottlenecks and competing selection pressures resulting in the appearance and disappearance of specific mutations.

### Impact of treatment and host immunity on BA.1.23 evolution

To determine the potential impact of SARS-CoV-2 antiviral treatments on intrahost evolution we examined the medication histories of patients P1 (index), P2, P3 and P4 (**Fig. 3**). We also assessed SARS-CoV-2 spike binding antibody levels using available serological data from clinical tests. The index patient P1 was vaccinated at the time of admission with two doses of an unspecified vaccine administered six months and one week before hospitalization. A third vaccine dose was administered two days after admission. Additional SARS-CoV-2 antiviral treatments included a course of remdesivir on days 1-6 and a dose of Gamunex IgG on day 9 (**Fig. 3D**). Moderate titers of SARS-CoV-2 antibodies were detected on day 38, around the time the first intra-host mutations were found (**Fig. 2C**). These titers could be due to residual antibodies from the Gamunex IgG treatment, an immune response to vaccination and/or infection, or a combination of treatment and host response. Notably, the detection of the first three spike mutations followed an increase in NAAT cycle thresholds (Ct) on day 30, suggesting a potential selection bottleneck around this time (**Fig. 2A**).

Patient P2 was fully vaccinated and had high titers of SARS-CoV-2 antibodies when assayed three months before and one month after their single positive SARS-CoV-2 NAAT (**Fig. 3B**). This patient also received a one-month course of nirmatrelvir/ritonavir (Paxlovid™) (**Fig. 3D**). The combination of high levels of host antibodies and antiviral treatment likely explains why this patient did not develop a persistent infection.

The long-term case P3 was fully vaccinated at the time of their first positive test and received a 3-week course of nirmatrelvir/ritonavir (Paxlovid™). However, no SARS-CoV-2 antibodies were detected in serological assays performed during the first 4 months of their persistent infection (**Fig. 3B**). Notably, during this time we also did not detect new substitutions in the BA.1.23 lineage, despite indications of active viral replication based on consistently low NAAT Ct values (less than 25) in NP and AN samples. Active replication was confirmed by isolation of replication-competent SARS-CoV-2 from five specimens collected between study days 101–147. Notably, the appearance of new intra-host substitutions during the last month of persistent infection occurred around the time of detection of SARS-CoV-2 antibodies at levels close to the lower limit of the serological assay (**Fig. 3**). P3 did not receive antibody therapy during their hospitalization, indicating that the seroconversion was the result of a delayed and weak host immune response.

Patient P4 was unvaccinated and received bebtelovimab mAb after their first positive SARS-CoV-2 PCR test, followed by a course of nirmatrelvir/ritonavir. P4 had low SARS-CoV-2 antibody titers at the time of the first positive PCR prior to the mAb treatment (**Fig. 3B**). The S:V445A mutation that emerged in this patient may be associated with the mAb treatment, as it was shown to result in a 111-fold reduction in susceptibility to bebtelovimab for viral variants carrying this mutation in pseudotyped virus-like particle (VLP) neutralization assays^21^. We did not find SARS-CoV-2 mutations associated with remdesivir^22-27^ or nirmatrelvir/ritonavir resistance^28^ in any of the patients treated with these drugs.

### Serum neutralization profile of Omicron BA.1.23

Neutralization of Omicron BA.1 isolates by sera from convalescent or twice boosted individuals is strongly reduced compared to the levels obtained for neutralization of the ancestral strains^29-31^. We therefore assessed the degree of neutralizing activity of BA.1.23 (BA.1 + S:E96D, R346T, L455W, K548M, E484V, P681R, A688V) compared to Omicron BA1 (B.1.1.529), and the ancestral SARS-CoV-2 (USA-WA1/2020, WA). We used a multi-cycle replication assay in which serum is present continuously to best mimic physiological *in vivo* conditions. We selected sera from two subsets of PARIS study participants representing distinct levels of immunity. The first panel of sera tested was collected before and after booster vaccination while the second set of samples was obtained before and after Omicron BA.1 breakthrough infection.

Prior to booster vaccination with monovalent SARS-CoV-2 RNA-based vaccines, neutralization of BA.1, BA.1.23 and the WA1/USA were comparable (geometric mean titer; GMT WA1: 31; GMT BA.1: 12; GMT BA.1.23: 9; **Fig. 4A**). However, 4/9 and 6/9 samples failed to display any neutralization activity against BA.1 and BA.1.23 respectively while this was true for only 2/9 samples when looking at activity against the ancestral virus. After booster vaccination, the loss in neutralization for BA.1.23 was greater than 15-fold compared to only a three-fold reduction for BA.1 relative to WA1/USA (geometric mean titer; GMT WA1: 674; BA.1:234; GMT BA.1.23: 45; **Fig. 4A**). Of note, the loss of neutralization activity for BA.1 measured here is less than what has previously been reported which could be due to assay variation in combination with the limited number of samples tested^29^.

**Figure 4.**
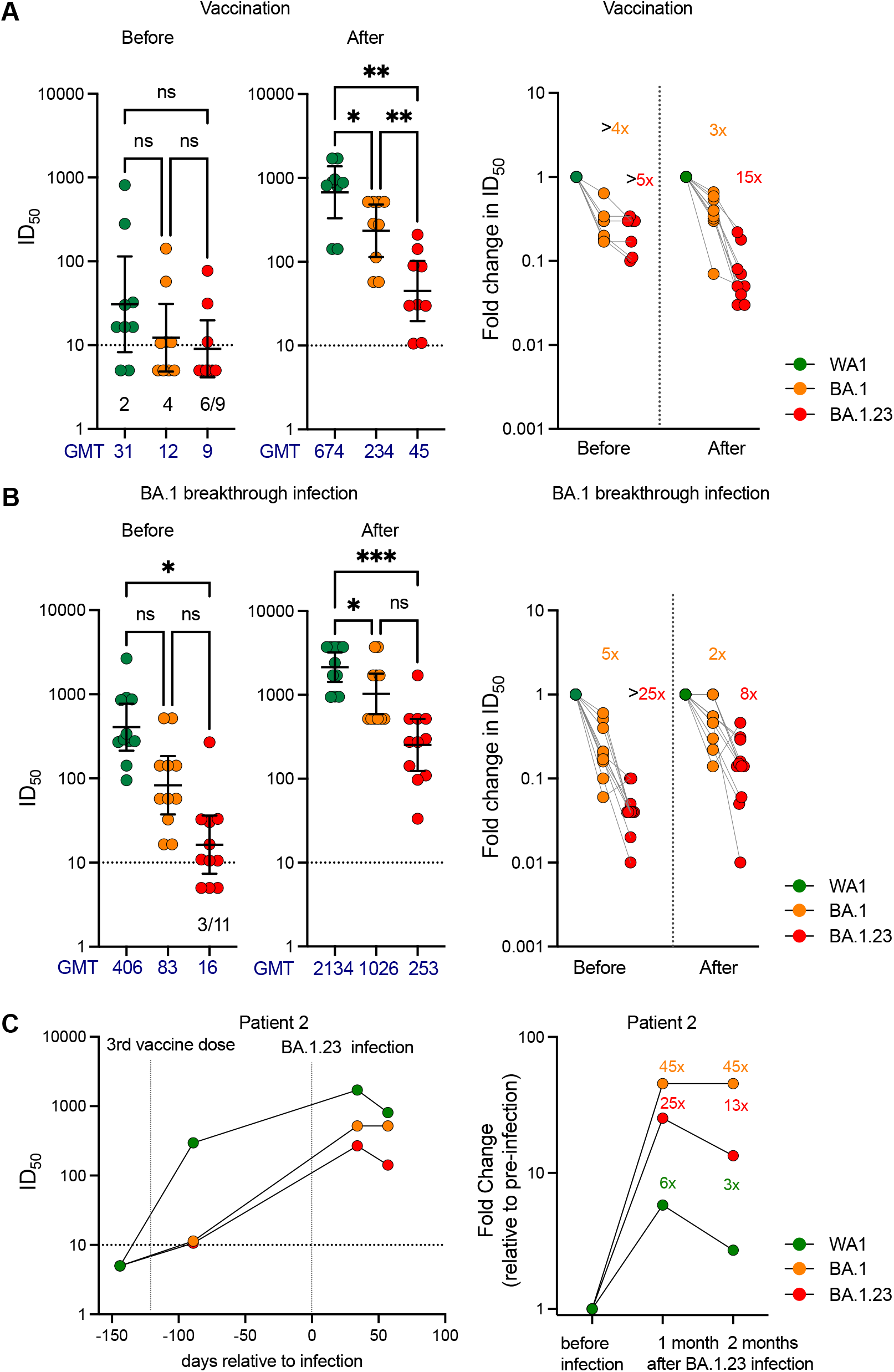
The Omicron BA.1.23 subvariant displays strongly reduced neutralizing activity compared to the parental Omicron variant as well as ancestral SARS-CoV-2 variant. **A)** Absolute neutralization titers (left) and fold reduction (right) for WA-1, BA.1 and BA.1.23 variants by paired sera from study participants collected before and after booster vaccination. The two samples with levels below detection for WA-1 were excluded from the fold-change calculation shown in the right panel. **B)** Absolute neutralization titers (left) and fold-reduction (right) for WA-1, BA.1 and BA.1.23 isolates by sera from study participants who experienced breakthrough infection with BA.1. Data for paired sera from 11 participants collected before and after BA.1 infection are shown. **C)** Absolute neutralization titers (left) and fold-changes in neutralization titers relative to the post booster timepoint (right) for each of the isolates (WA-1, BA.1 and BA.1.23) by sera collected from patient P2 before and after BA1.23 infection. The first vertical dotted line (left) represents the third vaccine dose. The second vertical dotted line (right) indicates the infection with the BA.1.23 variant. GMT denotes mean geometric mean of the inhibitory dilution 50% (ID50) values. The horizontal dotted line represents the limit of detection (10). Samples with neutralization titers below the level of detection were assigned the neutralization value of 5 (equaling to half of the limit of detection). Fold changes in neutralization are A one-way ANOVA with Tukey’s multiple comparison test was used to compare the neutralization titers before and after booster vaccination or BA.1 breakthrough infection. Significant p values (<0.05) are indicated in the figure. Source data is provided in **Supplementary Table S2**.

We next looked how sera collected before and after breakthrough infection would neutralize BA.1.23. We noted a significant difference in neutralization activity for BA.1.23 but not BA.1 compared to WA1 in the samples collected before and after BA.1 breakthrough infection (**Fig. 4B**). Prior to BA.1 infection, the difference was >25-fold with 3/11 samples having undetectable neutralization activity for BA.1.23 and 5-fold difference for BA.1 (GMT WA1: 406; GMT BA.1: 83; GMT BA.1.23: 16; **Fig. 4B**). After infection, neutralization titers increased for all three viruses reducing the difference to two-fold for BA.1 and 8-fold for BA.1.23 (GMT WA1: 2134; GMT BA.1: 1026; GMT BA.1.23: 253; **Fig. 4B**).

Lastly, we analyzed sera collected from patient P2 at different time points (e.g., before booster vaccination, after booster vaccination but before BA.1.23 infection, and at two time points after infection with BA.1.23). P2 mounted neutralizing antibodies after booster vaccination with the ancestral variant being neutralized >25 times more efficiently than BA.1 or BA.1.23 (**Fig. 4C)**. One month after breakthrough infection with BA.1.23, neutralization of all three viruses increased relative to the titers measured after booster vaccination but to very different degrees (e.g. WA1: 6-fold, BA.1:45 fold, BA.1.23: 25-fold). Of note, two months after infection, the BA.1 titers remained stable, while neutralization of the ancestral virus as well as BA.1.23 declined equally.

Collectively these results indicate that BA.1.23 is significantly more resistant to neutralization than the parental BA.1 variant.

## Discussion

It has been speculated that the emergence of antigenically diverse SARS-CoV-2 variants such as Omicron result from intrahost viral evolution driven initially by suboptimal immune responses and then spread by forward transmissions. Our data demonstrate that intrahost evolution of SARS-CoV-2 during persistent infection in immunocompromised individuals can drive the emergence and spread of novel (sub)variants with extensive mutations in key antigenic regions, even in the context of the already highly mutated Omicron lineage. Further sequential evolution in transmission cases shows that rapid divergence can occur stepwise and persist in small numbers of individuals, complicating early detection of novel variants.

The most extensive changes in the evolution of the BA.1.23 lineage were seen during its emergence in patient P1 and its subsequent divergence in patient P3 over a total period of 8 months. Based on available clinical testing data, both patients were initially negative for SARS-CoV-2 antibodies but seroconverted to moderate to low levels around the time spike mutations were accumulating. These conditions are likely optimal for the selection of immune escape mutations and provide an explanation for the clustering of BA.1.23-specific changes in the spike protein. It is important to note that the antiviral treatments administered over limited time periods as monotherapies (e.g., monoclonal antibodies, remdesivir, and nirmatrelvir/ritonavir (Paxlovid™) failed to eliminate the persistent infection, highlighting the need for improved or potential combination therapy tailored for these special situations.

During the initial emergence of the BA.1.23 lineage in index patient P1 mutations were concentrated in the spike protein. The amino acid substitutions or positions at which changes accumulated have been associated with neutralization escape (e.g., R346T, E484V, S477)^16,17^, increased angiotensin converting enzyme 2 (ACE2) binding avidity (e.g., R346T, L455W)^32^, improved viral fusogenicity (e.g., P681R/Y)^18^, or a predicted increase in fitness (e.g., positions 346, 484 and 681)^33^. Subsequent stepwise changes in patient P3 occurred more broadly throughout the SARS-CoV-2 genome. The RBD mutations N448S and F456L present in the most recent specimen from P3 are within the ACE2 binding region with potential for changes in binding affinity and antibody escape^32^. Mutations outside the spike mapped to nonstructural proteins, many of which have previously been detected in other VOCs such as Nsp3_T183I (Alpha), Nsp3_K977Q (Gamma), Nsp12_G671S (Delta AY.* and Omicron BA.2.75). Although the effects of these mutations have not been characterized, they occurred in the virus polymerase subunit (Nsp12) and in proteins with antagonistic activity towards the host’s innate immunity (Nsp3, ORF6, ORF7a, ORF9b)^34^.

Consistent with the extensive changes in the spike protein, resistance of the transmitted BA.1.23 variant to neutralization was substantially increased compared to the ancestral SARS-CoV-2 strain as well as to BA.1 Omicron. This was true both for sera collected before and after monovalent booster vaccination as well as after BA.1 breakthrough infection (**Figure 4**). Escape from neutralization was likely mediated by the combination of R346T, L455W, K458M, and E/A484V within the receptor binding region of spike. Of note, substitutions at position R346 are found in several Omicron sublineages (e.g., BA.4, BA.5) and subvariants (e.g., BA.2.72.2, BA.4.6/BF.7 (R346T), BA.4.7 (R346S), and BA.5.9 (R346I))^35^. These variants were not circulating at the time of BA.1.23 emergence pointing to convergent evolution in the context of suboptimal immune responses.

We further found that changes in minority viral populations can greatly increase the spectrum of mutations during persistent infection beyond what is observed at the consensus level. Viral ‘quasi-species’ have been described for pandemic coronaviruses such as SARS-CoV-1, Middle Eastern Respiratory Syndrome Coronavirus (MERS-CoV) and SARS-CoV-2^36-42^, but their role remains to be fully elucidated^43^. Interestingly, mSNVs predominantly occurred in conserved regions and remained present in multiple sequential specimens. Only a small subset of these mutations became dominant in later specimens. We speculate that these viral populations enable sub-optimal mutations to persist within the host, increasing the likelihood for the emergence of compensatory mutations that offset their fitness defects.

Despite careful monitoring, we did not encounter new BA.1.23 cases in our routine SARS-CoV-2 surveillance or in the GISAID database between June-September 2022, suggesting that the lineage has not spread further beyond the initial outbreak. This may be due to increased caution by immunocompromised patients to avoid contacts that could increase their risk of infection. We also note that patients P1, P3 and P4 were hospitalized for long stretches of time during their persistent infections, further limiting community exposure. Finally, the limited transmission could reflect a reduced fitness of BA.1.23 compared to contemporary lineages. Notable in this respect is that the emergence of the BA.1.23 lineage coincided with the general displacement of BA.1 by BA.2 sublineages in the NYC area, in particular BA.2.12.1, which dominated SARS-CoV-2 cases during the spring and early summer of 2022. A longer observation period with continued background surveillance will be needed to rapidly identify any potential re-emergence of BA.1.23 in patients other than the ones included in this study.

Our findings add to the accumulating knowledge regarding SARS-CoV-2 persistent infections, and document subsequent forward transmissions. Furthermore, we show that persistent infections can drive emergence of viral variants with the potential to spread to other individuals, some of whom may themselves develop prolonged infections, and thereby establish a pathway for continued virus evolution. This warrants genomic monitoring of persistent infections in particular, and further underscores the need to limit the duration of viral infection. Improved early detection of novel SARS-CoV-2 variants and forward transmission tracking, a better understanding of the selection processes driving SARS-CoV-2 evolution and the role of mSNVs, as well as therapies that limit virus persistence and shedding, are essential to reduce the emergence of highly mutated viral variants in the future.

## Materials and Methods

### Molecular SARS-CoV-2 diagnostics

SARS-CoV-2 molecular diagnostic testing was performed in the Molecular Microbiology Laboratories of the MSHS Clinical Laboratory by nucleic acid amplification tests (NAAT) that have been validated for nasopharyngeal (NP), anterior nares (AN) swabs, and saliva specimens. All but one positive sample included in this study were tested using the PerkinElmer® New Coronavirus Nucleic Acid Detection Kit which provides qualitative detection of nucleic acid from SARS-CoV-2. It includes two SARS-CoV-2 targets (ORF1ab, N) and one internal positive control (IC; bacteriophage MS2). Cycle threshold (Ct) values are generated for all three targets and provide a quantitative estimate of the viral load. The only other positive sample that was run on another testing platform was the first specimen of the index case (P1). It was tested as a point of care test (POC) using the cobas^**®**^ Liat^**®**^ System (cobas^**®**^ SARS-CoV-2 & Influenza A/B) assay, and the biospecimen was discarded prior to transfer to the MS-PSP.

### The Mount Sinai Pathogen Surveillance Program (MS-PSP)

The Mount Sinai Pathogen Surveillance Program (MS-PSP) has profiled the evolving landscape of SARS-CoV-2 in New York City (NYC) since the beginning of the pandemic^44,45^. We routinely perform complete viral genome sequencing of randomly selected contemporaneous SARS-CoV-2 positive specimens collected from patients seeking care at our health system. Residual respiratory specimens (NP, AN swabs) were collected after completion of the diagnostic process, as part of the Mount Sinai Pathogen Surveillance Program. The program was reviewed and approved by the Mount Sinai Hospital (MSH) Institutional Review Board (IRB-13-00981). The detailed investigation into persistent SARS-CoV-2 infections in patients receiving care at the Mount Sinai Health System was separately reviewed and approved by the MSH IRB (IRB-22-00760).

### Human Serum Samples

We used a panel of sera collected as part of the longitudinal observational PARIS (Protection Associated with Rapid Immunity to SARS-CoV-2) cohort^46^. This study follows health care workers longitudinally since April 2020. Samples were selected based on the study participants different levels of immunity (before and after booster vaccination, as well as before and after Omicron BA.1 infection). The study was reviewed and approved by the MSH IRB (IRB-20-03374). All participants signed written consent forms prior to sample and data collection and provided permission for sample banking and sharing. Sera collected before and after SARS-CoV-2 booster vaccination (N=9 participants, 18 sera) as well as before and after Omicron BA.1 break-through infections (N=11 participants, 22 sera) were selected for testing in this study (see **Supplementary Table S3** for infection, vaccine and demographics information).

In addition, patient P2 participated in another of our observational study (IRB-16-01215). They provided written consent prior to biospecimen and data collection. Several serum samples from this patient before and after BA.1.23 infection were available for neutralization assays.

The 44 sera (40 PARIS, 4 from P2) from our observational cohorts are unique to this study and are not publicly available.

### RNA extraction and SARS-CoV-2 genome sequencing

RNA was extracted using the Chemagic ™ Viral DNA/RNA 300 Kit H96 (PerkinElmer, CMG-1033-S) on the automated Chemagic ™ 360 instrument (PerkinElmer, 2024-0020) from 300uL of viral transport media per the manufacturer’s protocol. cDNA synthesis followed by whole genome amplification with two custom primer panel sets targeting 1.5 and 2kb regions across the SARS-CoV-2 genome was performed as previously described^44^ with the addition of new oligonucleotides to minimize amplicon dropout for Omicron lineages derived from PANGO lineage B.1.1.529 (**Supplementary Table S4**). Paired-end (2×150bp) Nextera XT (Illumina, cat. FC-131-1096) libraries prepared from amplicons were sequenced on a MiSeq instrument. SARS-CoV-2 genomes were assembled using a custom reference-based pipeline as previously described^44^. The final average sequencing depth per genome ranged between ∼135k to ∼415k reads.

### Analysis of minor nucleotide variants

Minority intra-host single nucleotide variants (mSNVs) were annotated when present in both reads of paired-end reads of a single sample and at a minimum frequency of 0.1 (10%). A second quality control filter was applied by only including positions for which mSNVs were present in more than one sample from the study (from different time pointes) or for positions that changed at the consensus level at any point of the investigation.

### Phylogenetic analysis

Global background SARS-CoV-2 sequences were downloaded from the GISAID EpiCOV database (as of 2022-08-30). The GISAID database was queried for the novel mutations observed in P1-P4 sequences to identify their closest matches. Sequence hits were confirmed by their Mash distance^47^. For this, a genome sketch was generated with Mash v.2.3 from the sanitized alignment of the global sequences filtered for lineages BA.1.* as produced by Nextstrain^1,48^ v11 for SARS-CoV-2 with default parameters (https://github.com/nextstrain/ncov). This allowed pairwise comparisons of our data with high-quality global sequences with available metadata. Maximum likelihood phylogenetic inferences of the MS-PSP SARS-CoV-2 genomes, including the closest sequence matches and all other BA.* lineage sequences from the MS-PSP surveillance program, were done using an Omicron BA.* and New York State-focused background with proximity subsampling scheme with Nextstrain under the GTR+G model of nucleotide substitution. Branch support was assessed with the ultrafast bootstrap method with 1000 replicates in IQ-TREE multicore version 2.1.2 [PMID: 29077904, 25371430]. Clade and lineage assignments were done with Nextclade CLI v3.2 (2021-11-04)^49^ and with PANGO-v1.8 (pangolin v3.1.17, pangoLEARN v.2022-04-22) ^50,51^.

### SARS-CoV-2 viral cultures

Replication-competent SARS-CoV-2 was obtained as previously described^29^. Briefly, Vero-E6 cells expressing transmembrane serine protease 2 (TMPRSS2) (BPS Biosciences, catalog (cat.) no. 78081) were cultured in Dulbecco’s modified Eagle medium containing 10% heat-inactivated fetal bovine serum and 1% minimum essential medium (MEM) Amino Acids Solution, supplemented with 100 U/ml penicillin, 100 μg/ml streptomycin, 100 μg/ml normocin, and 3 μg/ml puromycin. 200ul of viral transport media from the nasopharyngeal or anterior nares swab specimen was added to Vero-E6-TMPRSS2 cells in culture media supplemented with 2% heat-inactivated fetal bovine serum, 100g/ml normocin, and 0.5 μg/ml amphotericin B. Cultures were maintained for a maximum of 10 days. Culture supernatants were collected and clarified by centrifugation (3,739g for 5 min) upon the appearance of cytopathic effects. Viral cultures were successful for the initial specimens obtained from P2, P3, and P4, but failed for all P1 specimens.

### Titration and concentration of viral isolates

Viral stocks were sequence-verified and then titered using the 50% tissue culture infectious dose (TCID_50_) method on Vero-E6-TMPRSS2 cells. Based on sequencing verification, viral isolate from P3 (PV56567) was selected for further experiments. Since the initial viral titer of the expanded BA.1. 23 PV56567 viral stock was too low (4×10^2 TCID_50_/ml) to perform microneutralization assays, we concentrated the viral isolate 4x using Pierce™ Protein Concentrator PES, 100K MWCO (Thermo Scientific, Catalog number: 88537) columns as described by the manufacturer. Briefly, culture supernatant was collected upon appearance of a cytopathic effect (CPE), clarified by centrifugation (3,739g, 5min) and then concentrated. Titers after concentration were 1.27×10^5 TCID_50_/ml.

### Micro-neutralization assays with replication-competent SARS-CoV-2 isolates

Sera collected from three different groups of study participants were used to assess the neutralization of wild type (WA1), BA.1, and BA.1.23 SARS-CoV-2 isolates. The first panel of samples includes sera collected before and after mRNA SARS-CoV-2 booster vaccination (N: 9, PARIS cohort), the second panel includes sera collected before and after BA.1 Omicron break-through infection (N:11, PARIS cohort). The last series includes longitudinal samples from P2, the first of the forward transmission case, collected prior as well as after the infection with BA.1.23.

Sera from study participants were serially diluted (three-fold) from a starting dilution of 1:10 using infection media consisting of minimum essential media (MEM, Gibco) supplemented with 2 mM L-glutamine, 0.1% sodium bicarbonate (w/v, Gibco), 10 mM 4-(2-hydroxyethyl)-1-piperazineethanesulfonic acid (HEPES, Gibco), 100 U/ml penicillin, 100 μg/ml streptomycin (Gibco) and 0.2% bovine serum albumin (MP Biomedicals). Diluted sera were incubated for one hour with 10,000 TCID_50_ of the three different viruses at room temperature. After the incubation, 120 μl of the serum-virus mix were transferred to Vero-E6-TMPRSS2 (plated in 96 well plates the prior day) and incubated at 37°C, 5% CO_2_ for one hour. The serum-virus mix was removed and 100 μl/well of the diluted sera were added in addition to 100 μl/well of MEM 2% FBS. The plates were incubated for 48 hours at 37°C incubation. The cells were fixed (200 μl/well, 10% formaldehyde solution) overnight at 4°C prior to staining an anti-SARS nucleoprotein antibody produced in house (mAb 1C7C7)^52^. All procedures above were performed in the Biosafety Level 3 (BSL-3) facility at the Icahn School of Medicine at Mount Sinai following approved standard safety guidelines. The nucleoprotein staining was performed as previously described^29^. The analyses were performed using Prism 9 software (GraphPad).

### Statistics

Statistical analyses were performed using Prism 9 software (GraphPad). A one-way ANOVA with Tukey’s multiple comparisons test was used to compare the neutralization titers for the three viruses.

## Data Availability

All data produced in the present work are contained in the manuscript or available upon reasonable request to the authors

https://www.gisaid.org

## Acknowledgments

We thank the Mount Sinai Hospital and School Leadership, in particular Dr. David Reich, Dr. Dennis Charney, and Dr. Carlos Cordon-Cardo, for their ongoing support of the MS-PSP throughout the pandemic. We thank the laboratory technologists and staff in the Molecular Microbiology Laboratories and the Rapid Response Laboratories of the Mount Sinai Health System since without their assistance and help, none of this surveillance work would be possible.

We gratefully acknowledge the authors and originating and submitting laboratories of sequences from GISAID’s EpiCoV (www.gisaid.org) that were used as background for our phylogenetic inferences.

## Author contribution statement

Conceptualization: A.S.G.-R., H.A., E.M.S., V.S., H.v.B. Sample acquisition: S.S, G.P, J.P., A.A., A.R., C.C., D.F., L.S., C.G., K.S., J.D.R., R.B., P.S, A.P-M, PSP-PARIS Study Group. Sequencing and genome assembly: A.vd.G, K.F., Z.K., J.Z., B.A., R.S. Methodology: A.S.G.-R., H.A., N.A., J.M.C, F.K. Investigation: A.S.G.-R., H.A., J.M.C., E.M.S., V.S., H.v.B. Visualization: A.S.G.-R., H.A., V.S., H.v.B. Funding Acquisition: E.M.S., V.S., H.v.B. Project administration: K.S., E.M.S., V.S., H.v.B. Supervision: E.M.S., V.S., H.v.B. Writing – First draft: A.S.G.-R., H.A., H.v.B. Writing – Review and editing: A.S.G.-R., H.A., J.M.C, F.K. E.M.S., V.S., H.v.B.

## Additional information statement

Correspondence and requests for materials should be addressed to Drs. Sordillo, Simon and/or van Bakel.

## Data and code availability statement

Custom code used in this study is available at https://github.com/mjsull/COVID_pipe. Complete genome sequences for the viral isolates cultured from nasal swabs (BA.1 and BA.1.23) are available in GISAID.

## Extended Data Figures

**Extended Data Figure 1:**
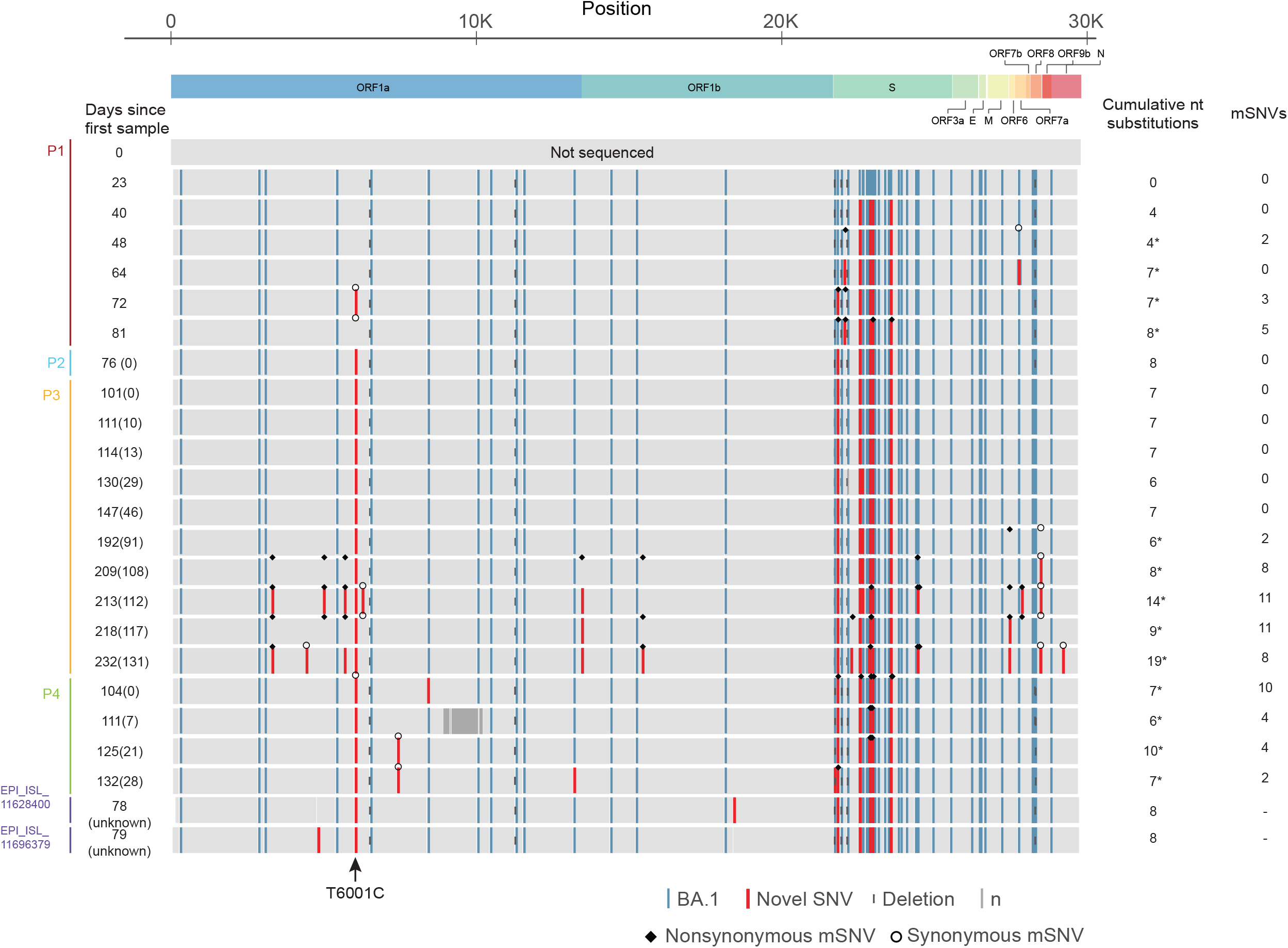
Overview of genome-wide SARS-CoV-2 mutations found in diagnostic nasal biospecimen. The mutations present across the complete SARS-CoV-2 genome are shown. Nucleotide substitutions observed in sequential specimens obtained from patient 1 (P1) with prolonged infection with BA.1 and forward transmission cases (P2, P3 and P4). There is accumulation and fixation of new SNVs in the spike region in P1. The same constellation of mutations was subsequently detected in three documented transmission cases (P2, P3 and P4) and in two sequences from GISAID (EPI_ISL_11628400 and EPI_ISL_11696379). The only shared synonymous SNV outside of the spike (ORF1a:T6001C) is indicated with an arrow. The number of days since the first positive test in P1 is shown on the left, with the number of days after the first positive test for each patient between brackets. The BA.1 substitutions are shown in blue and novel substitutions are shown in red, relative to the reference genome sequence NC_045512.2.

**Extended Data Figure 2:**
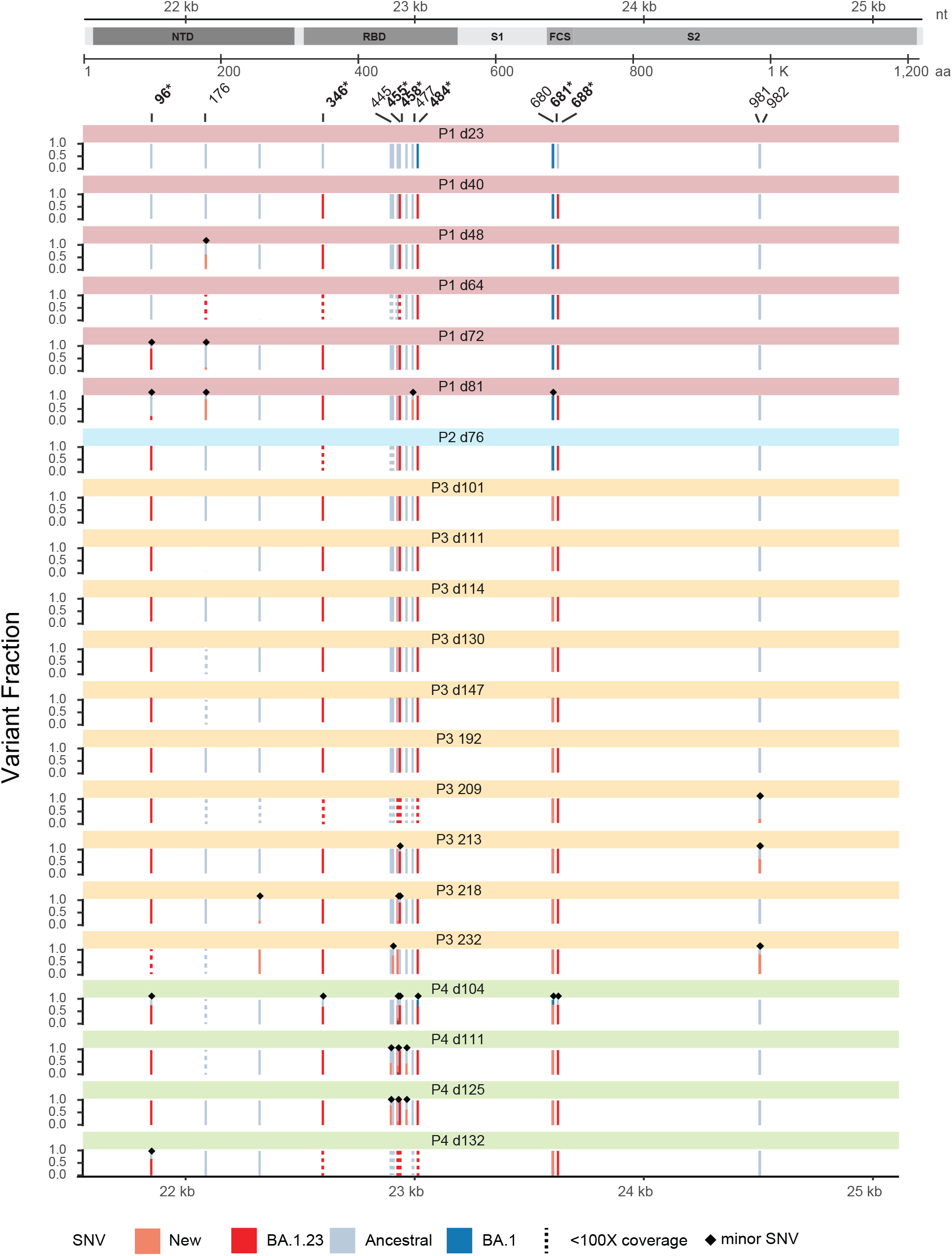
Summary of SARS-CoV-2 mutations within the spike gene in minority viral populations. The distribution of minority intrahost single nucleotide variants (mSNV) for positions that were fixed over the course of the infection of the index and transmitted infections are shown indicated with asterisks. Single nucleotide variants are colored by strain. The dotted bars indicate positions with coverage below the threshold of 100X coverage for calling mSNVs. The positions with minority variants present at frequencies > 0.1 are indicated with triangles for their respective samples.

**Extended Data Figure 3:**
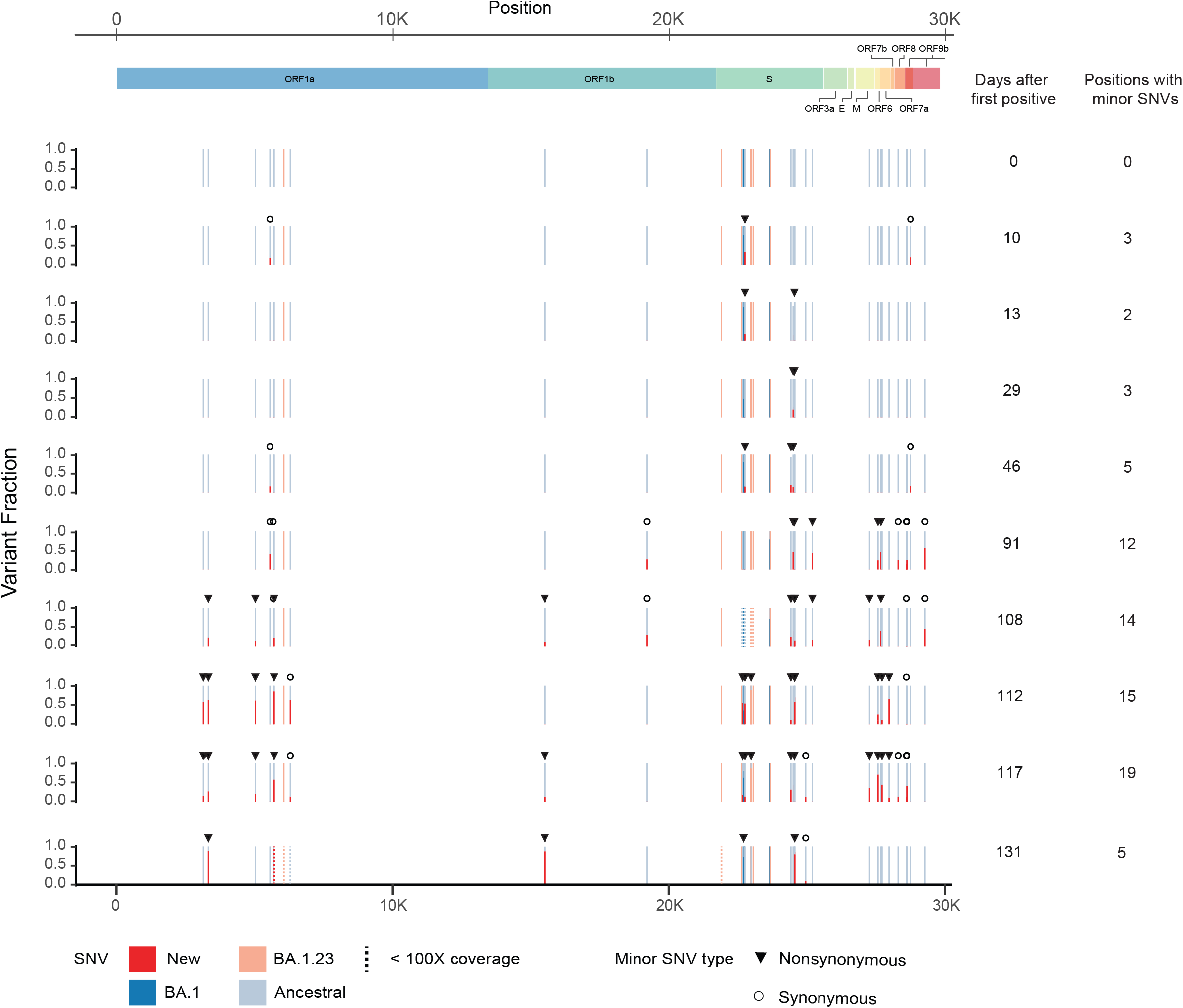
Summary of genome-wide SARS-CoV-2 mutations in minority viral populations for patient P3. Similar to Extended Data Fig. 2 but showing the distribution of nucleotide variants below the consensus level observed in one or more specimens from P3. Single nucleotide variants (SNVs) are colored by strain. The dotted bars indicate positions with coverage below the threshold of 100X coverage for calling mSNVs. The positions with minority variants present at frequencies > 0.1% are indicated with triangles (nonsynonymous) or circles (synonymous) for their respective samples.

**Extended Data Figure 4:**
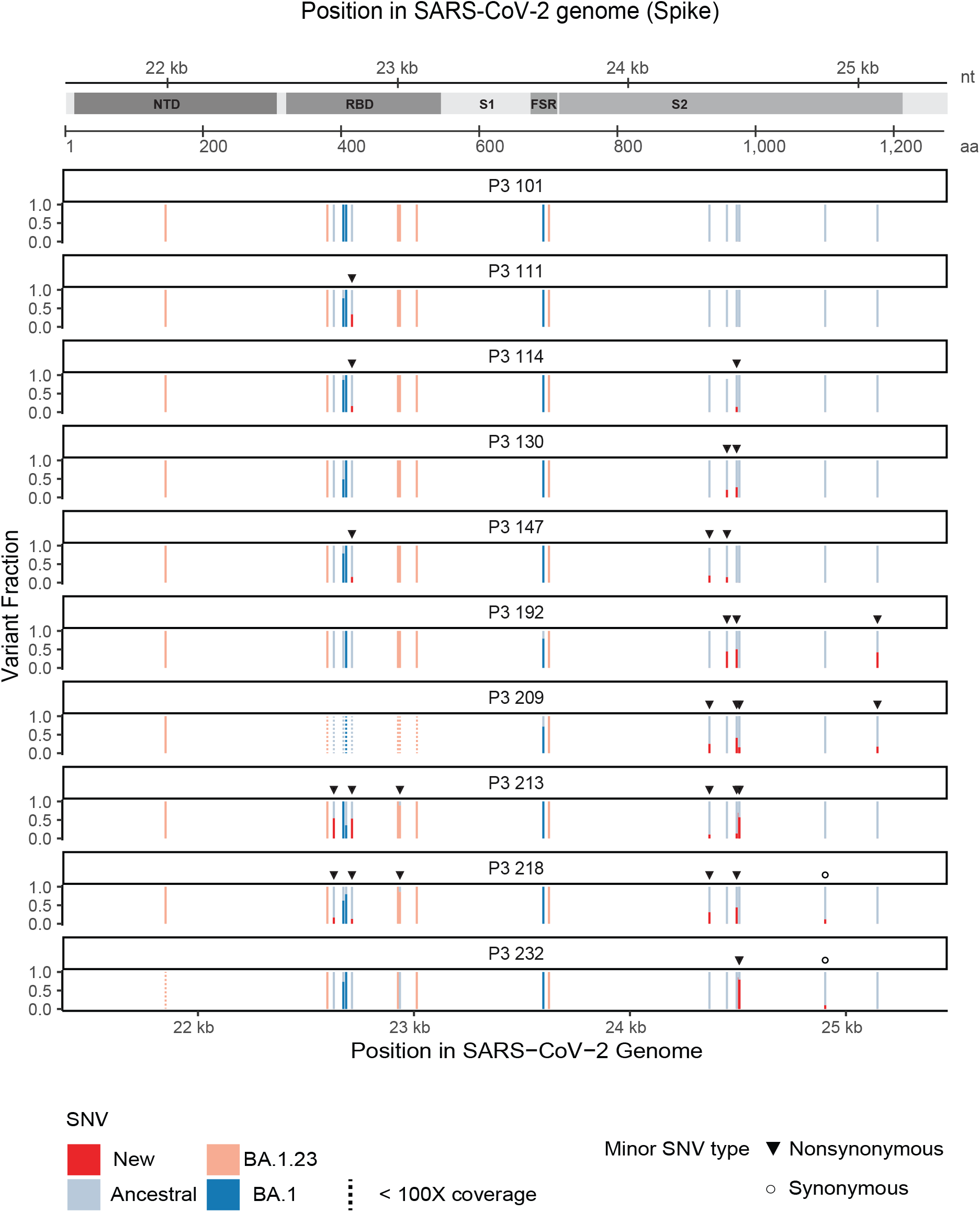
Summary of SARS-CoV-2 mutations within the Spike gene in minority viral populations for patient P3. Similar to Extended Data Fig. 3 but showing minority mutations in the Spike gene region. Single nucleotide variants (SNVs) are colored by strain. The dotted bars indicate positions with coverage below the threshold of 100X coverage for calling minor SNVs (mSNVs). The positions with minority variants present at frequencies > 0.1 are indicated with triangles for their respective samples.

